# Advancing CNS tumor diagnostics with expanded DNA methylation-based classification

**DOI:** 10.1101/2025.05.28.25328344

**Authors:** Martin Sill, Daniel Schrimpf, Areeba Patel, Dominik Sturm, Natalie Jäger, Philipp Sievers, Leonille Schweizer, Rouzbeh Banan, David Reuss, Abigail Suwala, Andrey Korshunov, Damian Stichel, Annika K Wefers, Ann-Christin Hau, Henning Boldt, Patrick N. Harter, Zied Abdullaev, Jamal Benhamida, Daniel Teichmann, Arend Koch, Jürgen Hench, Stephan Frank, Martin Hasselblatt, Sheila Mansouri, Theresita Díaz de Ståhl, Jonathan Serrano, Jonas Ecker, Florian Selt, Michael Taylor, Vijay Ramaswamy, Florence Cavalli, Anna S Berghoff, Brigitte Bison, Mirjam Blattner-Johnson, Ivo Buchhalter, Rolf Buslei, Gabriele Calaminus, Nicola Dikow, Hildegard Dohmen, Philipp Euskirchen, Gudrun Fleischhack, Amar Gajjar, Nicolas U Gerber, Marco Gessi, Gerrit H Gielen, Astrid Gnekow, Nicholas G Gottardo, Christine Haberler, Stefan Hamelmann, Volkmar Hans, Jordan R Hansford, Christian Hartmann, Frank L. Heppner, Pablo Hernaiz Driever, Katja von Hoff, Ulrich W Thomale, Stephan Tippelt, Michael C Frühwald, Christof M Kramm, Ulrich Schüller, Jens Schittenhelm, Martin U Schuhmann, Marco Stein, Petra Ketteler, Marc Ladanyi, Nada Jabado, Barbara C Jones, Chris Jones, Matthias A Karajannis, Ralf Ketter, Patricia Kohlhof, Uwe Kordes, Annekathrin Reinhardt, Christian Kölsche, Katrin Lamszus, Peter Lichter, Sybren L N Maas, Christian Mawrin, Till Milde, Michel Mittelbronn, Camelia-Maria Monoranu, Wolf Mueller, Martin Mynarek, Paul A Northcott, Kristian W Pajtler, Werner Paulus, Arie Perry, Ingmar Blümcke, Karl H Plate, Michael Platten, Matthias Preusser, Torsten Pietsch, Marco Prinz, Guido Reifenberger, Bjarne W Kristensen, Marcel Kool, Volker Hovestadt, David W Ellison, Thomas S Jacques, Pascale Varlet, Nima Etminan, Till Acker, Michael Weller, Christine L White, Olaf Witt, Christel Herold-Mende, Jürgen Debus, Sandro Krieg, Wolfgang Wick, Matija Snuderl, Ken Aldape, Sebastian Brandner, Cynthia Hawkins, Craig Horbinski, Christian Thomas, Pieter Wesseling, Andreas von Deimling, David Capper, Stefan M Pfister, David TW Jones, Felix Sahm

## Abstract

DNA methylation-based classification is integral to contemporary neuro-oncological diagnostics, as highlighted by the current World Health Organization (WHO) classification of central nervous system (CNS) tumors. We introduce the Heidelberg CNS Tumor Methylation Classifier version 12.8 (v12.8), trained using 7,495 methylation profiles, thereby expanding recognized tumor types from 91 classes in the previously published v11 ^1^ to 184 subclasses in v12.8. This expansion was primarily driven by novel tumor types discovered in our large website-derived repository and through global collaborations, further elucidating the heterogeneity of CNS tumors. Utilizing a random forest-based methodology, the classifier was rigorously validated through five-fold nested cross-validation, achieving a 95% subclass-level accuracy and a Brier score of 0.028, indicative of well-calibrated probability estimates. The hierarchical output structure facilitates comprehensive interpretation, allowing clinicians to assess subclass and aggregate class-level probabilities for informed decision-making. Comparative analyses demonstrate that v12.8 surpasses previous versions as well as traditional WHO-based diagnostics across diverse tumor cohorts. These advancements underscore the enhanced precision and practical utility of the updated Heidelberg CNS Tumor Methylation Classifier, reinforcing the pivotal role of DNA methylation profiling in personalized neuro-oncological care.

## Introduction

DNA methylation-based classification has become a central pillar of state-of-the-art diagnostics in neuro-oncology. Most prominently, the fifth edition of the World Health Organization classification of central nervous system tumors (CNS5, published in 2021)^2^ lists DNA methylation profiling as a desirable or even essential method for accurately diagnosing several tumor types. Parallel to these developments, other guideline authorities and medical societies, such as the National Comprehensive Cancer Network (NCCN)^3^, European Association of Neuro-Oncology (EANO)^4^, International Collaboration on Cancer Reporting (ICCR)^5^ or Royal College of Pathologists (RCPath UK), have also integrated methylation profiling into their recommendations. The concept of DNA methylation-based classification was introduced in 2016 with the initial public release of the Heidelberg CNS Tumor Classifier (v11), which was trained on data from a reference set later published by Capper et al. in 2018^1^. The classifier has been publicly available for the last nine years - free of charge - at molecularneuropathology.org, with an end user license agreement that allows users to opt in or out of sharing their data. This setup has facilitated large-scale accumulation of DNA methylation profiles from around the world, with over 160,000 sample data uploaded until the data freeze in October 2024 (Figure 1 a-b). The classifier has been utilized and validated in independent cohorts across diverse clinical settings, further demonstrating its robustness and clinical utility^6^. As the data repository continues to expand, it has offered new perspectives on the diverse landscape of brain tumors. Notably, a considerable number of uploaded samples failed to align with any of the 91 methylation classes in the original classifier (v11), prompting exploratory analyses that exposed previously unclassifiable tumor groups (Figure 1c). These findings laid the groundwork for creating an updated training dataset, encompassing a broader spectrum of methylation-defined or -supported entities (Figure 1d).

**Figure 1.**
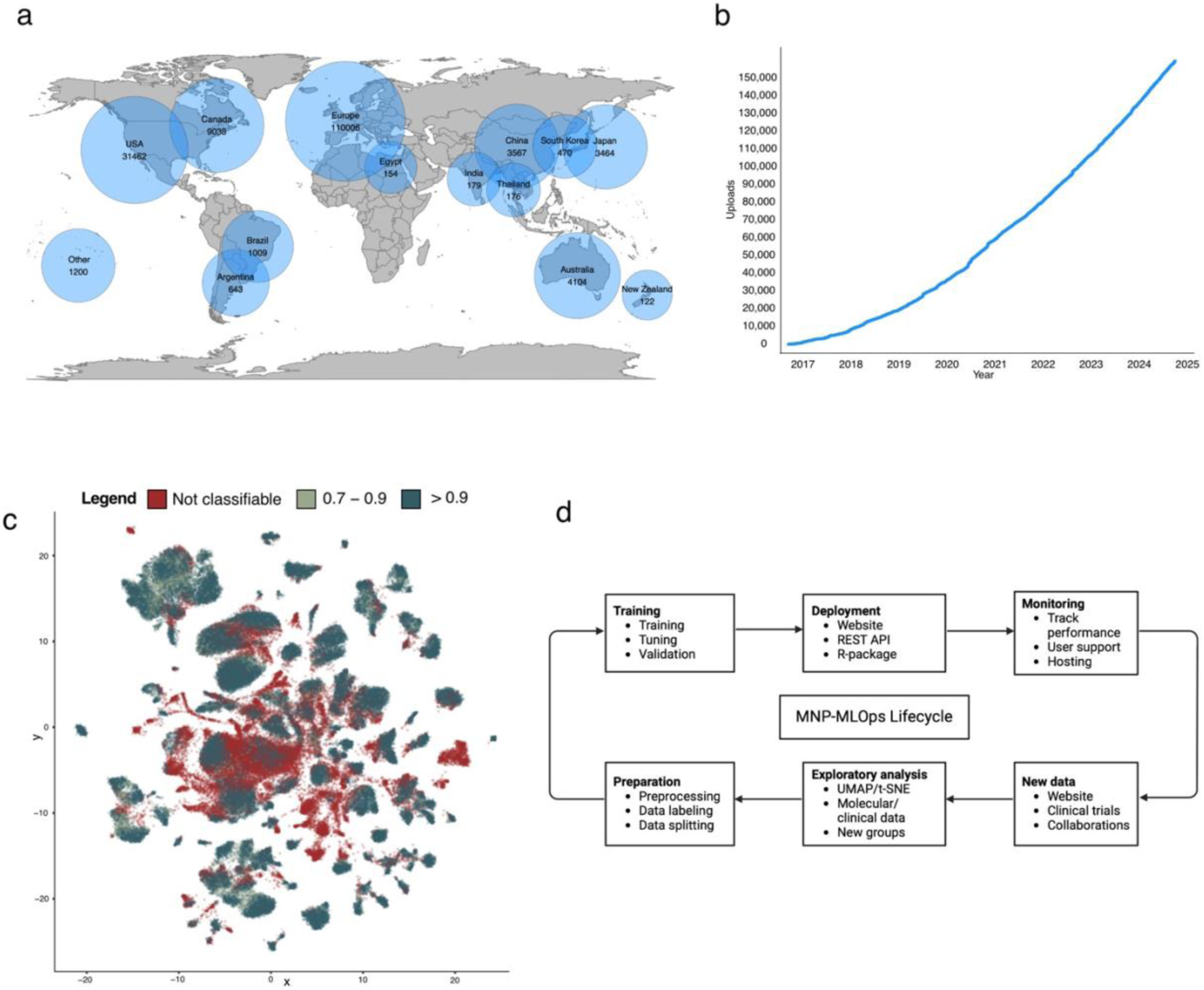
**Overview of the molecularneuropathology.org platform and its utilization** (a) Global distribution of sample uploads to molecularneuropathology.org from October 2016 to February 2025. Circle sizes on the world map are proportional to log(number of uploads) per country/region. (b) Temporal trend of sample uploads, showcasing the cumulative growth from 2016 to 2024. (c) UMAP projection of 97,213 CNS tumor samples, dots are colored by the v11 classifier scores for each sample (d) Flow diagram illustrating the Machine Learning Operations (MLOps) lifecycle used for model training, validation, deployment, and maintenance for the MNP classifier.

## Results

*Training Data Expansion, Exploratory Data Analyses, and Hierarchical Structure* Building upon the previously published Heidelberg Classifier v11 reference dataset comprising 2,801 samples, we expanded the training set to encompass 7,495 CNS methylation profiles (Figure 2). Of these, approximately 19% derive from the previous v11 cohort, preserving continuity and consistency with established diagnostic categories. An additional 11% originate from user submissions *via* our publicly accessible website platform. While this subset represents a relatively small proportion of the final training dataset, the samples uploaded to the platform played a valuable supporting role in identifying new tumor groups. The remaining samples were either diagnostic cases added to increase the sample size of previously underrepresented classes or sourced from institutional collaborations, particularly those focusing on well-characterized entities such as meningiomas^7^, posterior fossa (PF) A and PFB ependymomas^8,9^ and medulloblastomas^10,11^.

**Figure 2.**
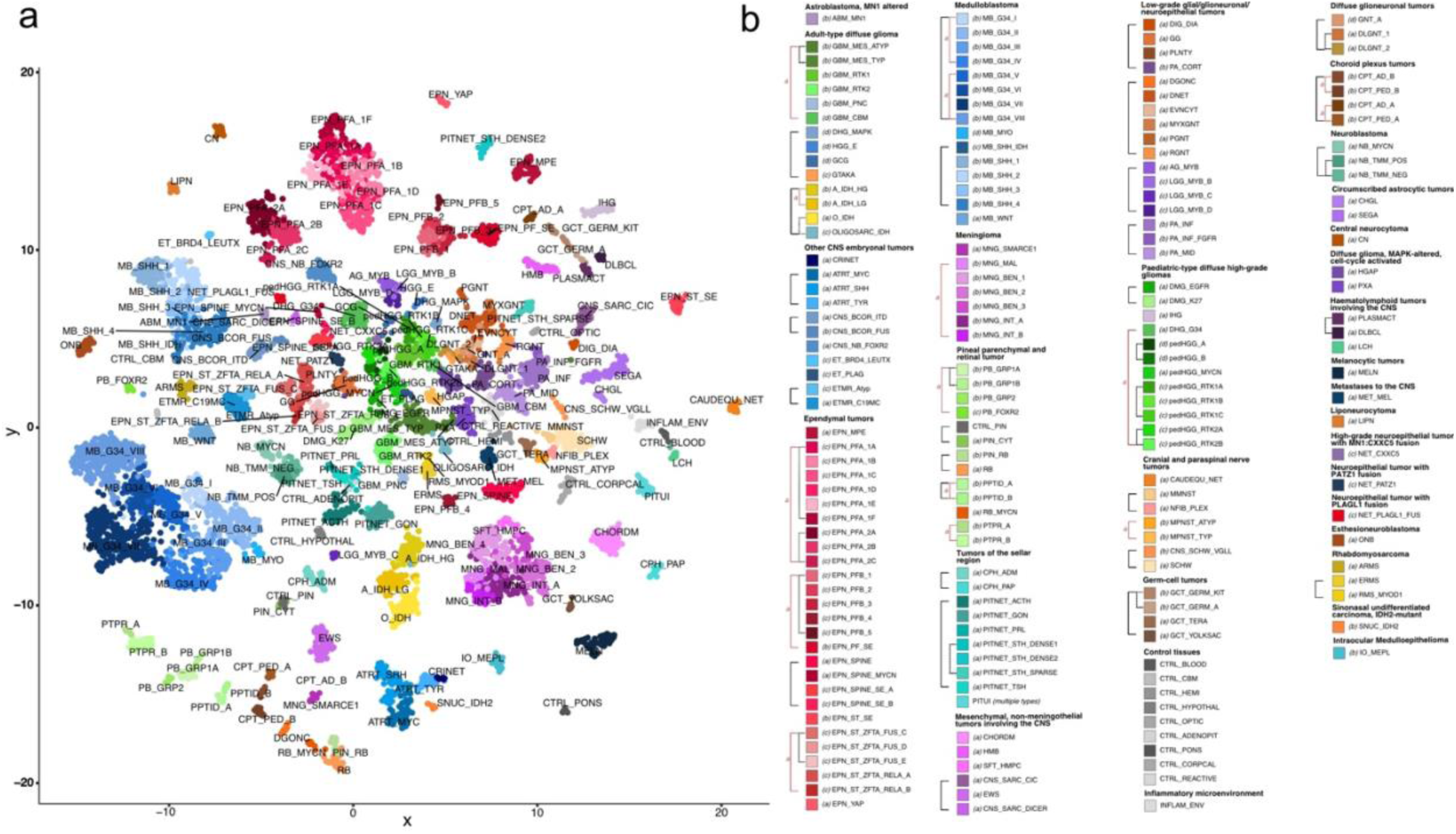
**Training dataset for v12.8** (a) UMAP projection of 7,495 samples used for training the v12.8 classifier (b) Legend indicating color code for each subclass shown in (a). Letters in rounded brackets before abbreviation of the subclass indicate ‘evidence level’ for the respective subclass. Each broad category shown in bold in the legend is a superfamily, families are indicated using a solid bracket on the second level adjacent to the colored blocks, classes are indicated using a dotted-line bracket at the first level. Red brackets indicate evidence level ‘a’ for the indicated class or family level

To leverage the growing volume of methylation profiles, we regularly performed non-linear dimensionality reduction analyses - specifically, t-Distributed Stochastic Neighbor Embedding (t-SNE) and Uniform Manifold Approximation and Projection (UMAP). These methods enabled unsupervised exploratory data analysis, revealing new, distinct clusters of samples that were previously unclassifiable under the original (v11) framework (Figure 3). Visual inspection of t-SNE and UMAP plots helped differentiate newly emerging tumor clusters from known entities, prompting us to define additional subtypes and refine existing ones. By systematically integrating these newly identified groups into a broadened classification scheme, the total number of recognized classes has effectively doubled from 91 to 184.

**Figure 3.**
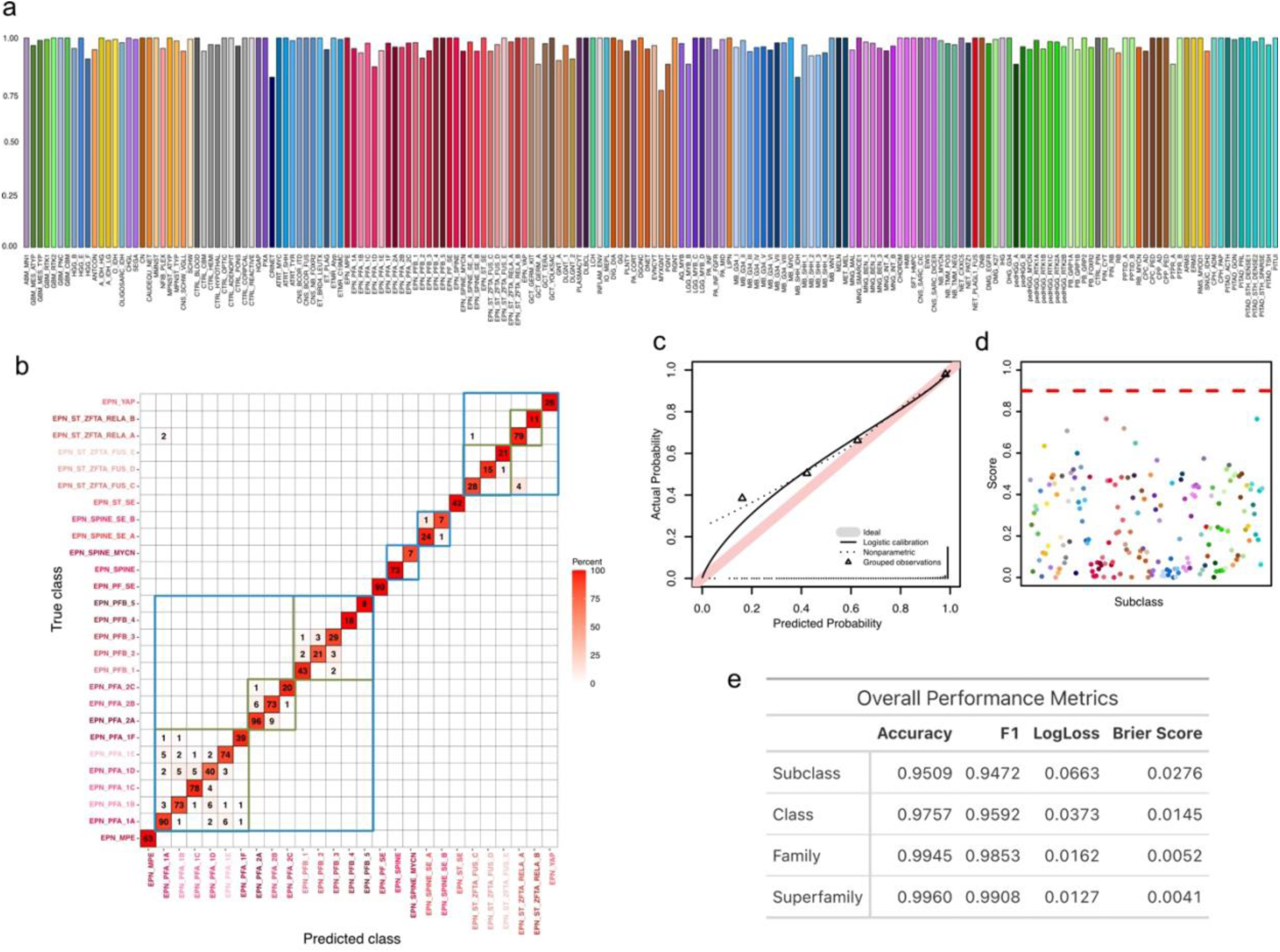
**Five-fold cross-validation performance of the v12.8 classifier** (a) Bar plot showing the balanced accuracy for each of the 184 subclasses, as derived from five-fold nested cross-validation. (b) Confusion matrix focusing on the ependymoma superfamily, where the majority of misclassifications occur between subclasses that belong to the same class or family (indicated by green and blue rectangles, respectively). (c) Calibration plot comparing predicted probabilities with observed outcomes, illustrating the degree of score calibration across subclasses. (d) F Scatterplot of subclass-specific Youden-optimal thresholds with color-coded tumor classes and a red dashed line at 0.9 marking the recommended threshold. (e) Table summarizing overall performance metrics—accuracy, F1-score, log loss, and Brier score—evaluated at each hierarchical level (subclass, class, family, superfamily).

These new groups encompass a diverse range of tumor classes and subclasses, including the well-established classes of meningiomas^7^, ependymomas subtypes PFA and PFB^8,9^, medulloblastomas^10,11^ and ependymomas characterized by different *ZFTA* gene fusions (previously known as RELA ependymomas)^12^, as well as newly added spinal ependymomas that show MYCN amplifications^13^, subtypes of diffuse leptomeningeal glioneuronal tumours (DLGNT) ^14^, diffuse glioneuronal tumors with oligodendroglioma-like features and nuclear clusters (DGONC)^15^, IDH-mutant oligosarcomas^16^, clear cell meningiomas with SMARCE1 mutations^17^, cribriform neuroepithelial tumors^18^, glioblastomas with primitive neuronal component^19^, glioneuronal tumor with ATRX alteration (GTAKA)^20^, subtypes of *MYB/MYBL1*-altered gliomas^21,22^, subtypes of previously unrecognized neuroepithelial tumors ^23–25^, malignant melanotic nerve sheath tumors ^26^, plexiform neurofibromas^27^, tumors with *EP300:BCOR(L1)* fusions ^28^, FOXR2-altered CNS neuroblastoma^29^, embryonal tumors with *BRD4::LEUTX* fusion ^30^, embryonal tumors with PLAG-family amplification^31^, germ cell tumors ^32–34^, subtypes of rhabdomyosarcoma^35,36^, sinonasal undifferentiated carcinoma *IDH2*-mutant^37^, intraocular medulloepithelioma^38^, Langerhans cell histiocytosis ^39^, subtypes of pineal parenchymal tumors of intermediate differentiation ^40^, neuroblastomas^41^, and MYCN-activated retinoblastoma ^42^.

Alongside this expansion, we introduced a four-tier hierarchical structure comprising superfamilies, families, classes, and subclasses (Figure 2). Established diagnostic categories generally reside at the family or class level, where sufficient clinical, histological and molecular evidence supports robust demarcation. By contrast, newly recognized groups - sometimes defined by rather subtle epigenetic variations - are often placed at the subclass level, acknowledging unknown clinical relevance at this early stage. In cases where the boundaries among subclasses remain poorly defined, the classifier defaults to higher-level group assignments as a conservative approach. This hierarchical organization thus accommodates both well-characterized tumor subtypes and newly discovered clusters, ultimately offering a more nuanced classification system that better captures the current knowledge and complexity of the CNS tumor landscape. To provide guidance on the available information on the new and existing classes and subclasses and their alignment with the current WHO classification, we introduced an evidence level annotation as specified in Figure 2 and Extended Data Table 1, curated and reviewed among an international group of neuropathologists: Level a - Tumor type/subtype identical to WHO 2021; Level b - Large single or more than one smaller dataset published describing the type/subtype as molecularly and/or clinically distinct, or the methylation class represents a distinct fraction of an established WHO 2021 tumor class; Level c - Single small dataset or case series; Level d - solely based on clusters in tSNE/UMAP. The annotation is typically provided at the most granular layer, plus a higher layer if the latter matches a WHO type/subtype.

### Classifier Training, Internal Validation and Comparison to v11

To train the classification model, we followed the Random Forest–based approach described in Capper et al. (2018)^1^ and Maros et al. (2020)^43^. To evaluate its performance, the classifier was validated using a five-fold nested cross-validation scheme. All subclasses achieved a balanced accuracy greater than 0.75, with 175 out of 184 subclasses exceeding 0.9 in accuracy. (Figure 3a). Overall, the classifier achieved a 95% subclass-level accuracy and a Brier Score of 0.028. In multiclass classification, the Brier Score measures the mean squared difference between predicted probabilities (that is, calibrated classifier scores) and observed class frequencies, indicating that the probability estimates are exceptionally well-calibrated and outperform those of the original v11 classifier (Figure 3b-d).

In addition, our newly introduced hierarchical system leverages these well-calibrated probabilities by aggregating subclass-level scores at higher tiers, thereby providing more robust diagnostic guidance at these higher-order tiers. As observed with the v11 classifier, most ‘errors’ occur between closely related subclasses or classes, such as among different subclasses (*) of Posterior Fossa Group A (EPN_PFA_*) ependymomas, between subclasses of group 3 and 4 medulloblastomas (MB_G34_*), or among benign meningioma subclasses (MNG_BEN_*) (Figure 3e). While some of these newly delineated subclasses indeed correlate with different prognostic outcomes, as described in their respective publications, most subclasses currently lack direct clinical implications, making aggregated probability scores at higher hierarchical levels sufficient for guiding diagnostic decisions according to current knowledge.

Among the samples in our methylation database, 97,143 achieved a v12.8 classifier score of ≥0.7 at the subclass level (Figure 4a). When applying the v11 classifier to this cohort, only 79,749 samples (82%) could be classified with a confidence score of ≥0.7 (Figure 4b-c). These additional classifiable cases included not only newly added subclasses but also some existing classes that now benefit from the increased training data available in v12.8. Of the samples unclassifiable in v11 (score <0.7), 2,128 (12%) were classified as Glioblastoma, IDH-wildtype, Mesenchymal Type, 1,422 (8%) as Glioblastoma, IDH-wildtype, RTK1/RTK2, and 587 (3%) as IDH-mutant Astrocytoma with v12.8 scores ≥0.7, thus underscoring the v12.8 classifier’s higher confidence in entities already established in v11. Overall, these results demonstrate the enhanced performance of v12.8, which not only accommodates newly discovered tumor types but also provides more robust classification for previously recognized entities.

**Figure 4.**
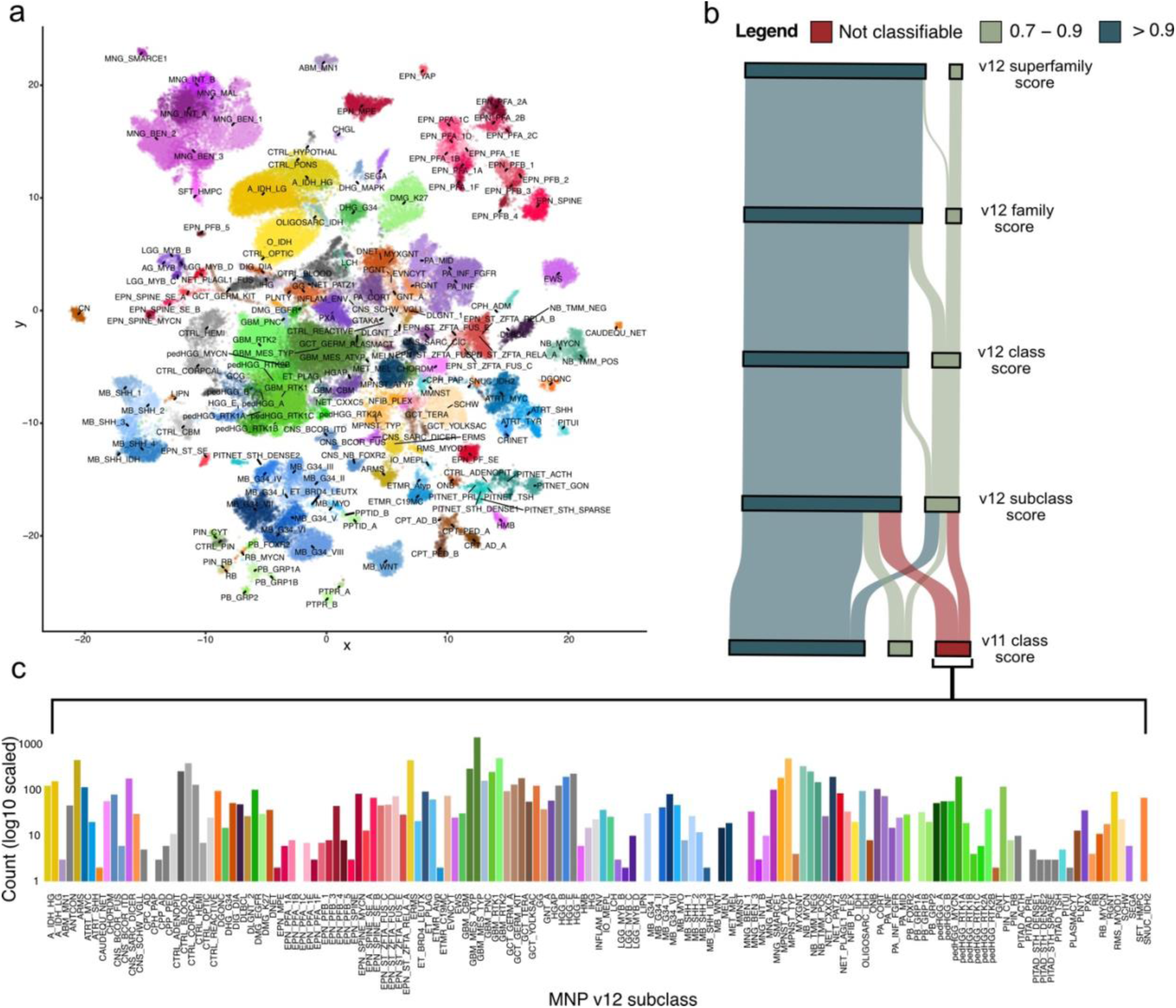
**UMAP projection comparing classification performance of mnp_v11b6 and mnp_v12.8** (a) Methylation profiles of 97,213 CNS tumor samples, including the v12.8 training data, classified using the v12.8 classifier, where all samples achieve a classification score of ≥0.7 and are colored according to the v12.8 color scheme. (b) Sankey plot showing scores for predictions with the v11 classifier and hierarchical levels of the v12.8 classifier respectively for the samples illustrated in (a). (c) Barplot indicates log10 scaled counts of v12 subclass predictions for samples not classifiable using v11.

### External Validation

To show the clinical potential of the v12.8 classifier, we analyzed data from the prospective, population-based Molecular Neuropathology 2.0 (MNP 2.0) study^44^, conducted within the German pediatric neurooncology ‘Treatment Network HIT’, which featured blinded central neuropathological review alongside molecular testing. In this cohort of over 1,200 newly diagnosed pediatric CNS tumor patients, the combined application of DNA methylation profiling and targeted panel sequencing improved the accuracy of tumor classification and identified cases where molecular data clarified ambiguous histology. Kaplan–Meier analysis of 80 ependymoma cases and 171 medulloblastoma cases, grouped by their v12.8 methylation subclass, revealed distinct survival curves for each subclass (Fig. 5a-b), highlighting meaningful prognostic differences. One important finding of the MNP2.0 study was that a subset of tumors originally diagnosed as high-grade gliomas (HGG) by conventional histopathology were classified as low-grade gliomas (LGG) when methylation results were taken into account; these patients showed more favorable outcomes during a median follow-up of 2.5 years, indicating that methylation-based classification could guide less intensive therapy. In line with this finding, Figure 5c shows a Sankey plot of 96 cases initially reported as ’high-grade gliomas’. Of these, 22% were assigned to a lower grade tumor subclass with a score of at least 0.9 by the v12.8 classifier.

**Figure 5.**
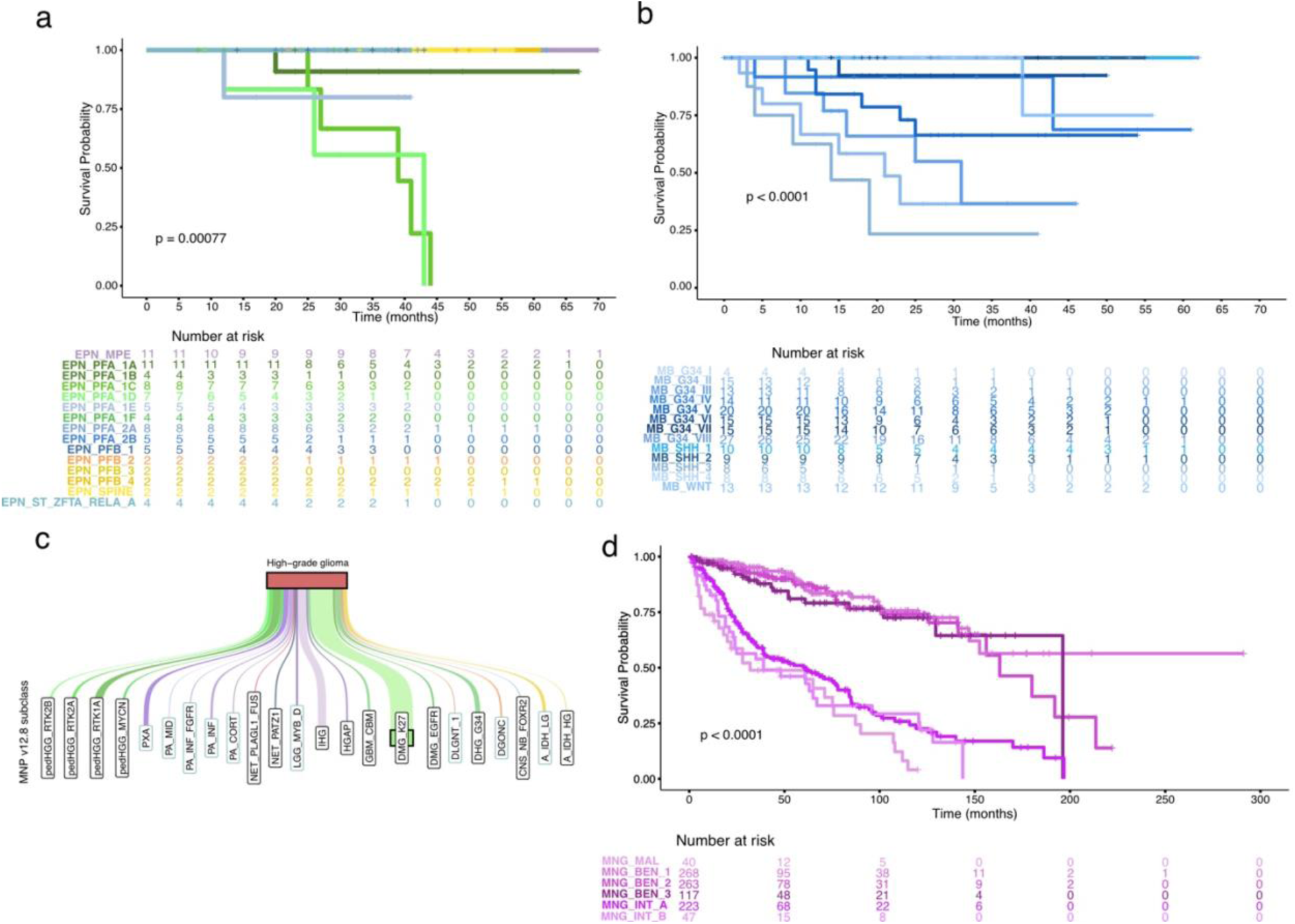
**Prognostic performance of the v12.8 classifier compared to alternative explanatory variables in Cox proportional hazards models.** (a) Kaplan–Meier estimates of overall survival for ependymoma patients from the MNP2.0 cohort stratified by v12.8 methylation subclass. The x-axis represents time in months, and the y-axis represents the survival probability. Differences between subclasses were assessed using the log-rank test; p < 0.05 was considered statistically significant. (b) Kaplan–Meier estimates of overall survival for medulloblastoma patients from the MNP2.0 cohort stratified by v12.8 methylation subclass. The x-axis represents time in months, and the y-axis represents the survival probability. Differences between subclasses were assessed using the log-rank test; p < 0.05 was considered statistically significant. (c) Sankey plot showing methylation subclass predictions (score >0.9) for histologically assigned high-grade gliomas in the MNP2.0 cohort. Subclasses outlined in blue indicate low grade tumours. (d) Kaplan–Meier estimates of progression-free survival from the meningioma Maas et. al 2021 cohort stratified by v12.8 methylation subclass. The x-axis represents time in months, and the y-axis represents the survival probability. Differences between subclasses were assessed using the log-rank test; p < 0.05 was considered statistically significant. Color of curves indicates v12.8 subclass.

In another large study of meningiomas, the DNA-methylation family assignment was combined with WHO histological grading and chromosomal copy number variation (CNV) data to develop an integrated molecular-morphologic score (intS) for risk stratification that significantly outperformed WHO grading alone ^45^. Hielscher et al. 2023^46^ investigated the intS and its clinical implementation, pointing out that using methylation classes or subclasses of meningiomas might improve the intS even more. To illustrate this potential of v12.8 methylation subclasses for risk stratification of meningiomas, Figure 5d shows the distinct Kaplan-Meier curves of predicted methylation subclasses for 958 meningioma patients included in this cohort.

Overall, these three studies exemplify that DNA methylation-based profiling allows for more accurate classification and can be used for better risk stratification of patients.

### Threshold Selection for Multiclass Classification

As with the previous v11 classifier, we recommend a 0.9 threshold across all tumor entities at the family level in v12.8. This threshold was chosen not only because it is straightforward to communicate but also because it has performed reliably in practical settings. We performed one-vs-all ROC analyses for each subclass and selected the threshold that maximizes Youden’s J, thus optimally balancing sensitivity and specificity. The highest Youden-based threshold was 0.77, which indicates that using a 0.9 threshold-even at the family level - is somewhat conservative. However, a 0.9 threshold helps ensure high specificity for certain classes. Ultimately, the well-calibrated probability estimates provided by our classifier allow users to interpret predictions in the context of other available clinical, histological, and molecular data.^47^

## Discussion

Identification of novel clusters, designation as subclasses and classes, and their subsequent implementation into diagnostic guidelines is inherently an iterative process. The recent work of the cIMPACT-NOW consortium^48^, endorsed by the International Society of Neuropathology, offers a framework for systematically evaluating emerging signals that suggest putative new tumor entities. Their guidelines recommend gathering comprehensive molecular, histopathological, and clinical outcome data before classifying any newly identified epigenetic cluster as a distinct diagnostic entity. The expanded Heidelberg Methylation Classifier (v12.8) exemplifies this iterative approach, wherein putative novel subgroups emerging from exploratory t-SNE and UMAP clustering undergo continuous scrutiny, validation, and consensus-building among international neuro-oncological experts.

Although conceived primarily as a research tool, the Heidelberg Methylation Classifier has, over time, demonstrated profound diagnostic potential, in part reflected by its inclusion in multiple neuro-oncology guidelines. Yet, the tension between rapid scientific progress and the rigor demanded by clinical regulatory environments poses challenges. For instance, national regulations initially restricted both the scope of distribution and the permitted application of the classifier website. Nonetheless, the growing evidence base and the recognized clinical impact of methylation-based classification spurred a shift toward broader diagnostic usage. In response to this demand, a commercial spin-off from the originating host institutions in Heidelberg has now been licensed to distribute local copies of the classifier, subject to applicable certification and reimbursement pathways. This model facilitates clinical adoption but also underscores the important role that regulatory frameworks play in ensuring safety and efficacy - particularly given that decisions about tissue-based diagnoses can directly dictate treatment plans. For research purposes, however, the classifier will still be made available free of charge, ensuring continued accessibility for academic and exploratory purposes.

The v12.8 expansion, alongside an enhanced hierarchical structure, reflects not only new entities but also the importance of shared terminology. Historically, subtle discrepancies existed between formal guidelines and the labels output by the classifier. These discrepancies partly stem from the fact that diagnostic guidelines evolve on the basis of established clinical evidence, whereas classifier outputs may temporarily adopt more provisional (and often more granular) subclass designations when new molecular subgroups first emerge. To bridge this gap, a global panel of neuropathologists and molecular neuro-oncologists convened to refine and align subclass annotations, culminating in the updated nomenclature for both newly discovered and longstanding tumor classes applied here. Such efforts ensure that the classifier keeps pace with refinements in disease knowledge while maintaining coherence with diagnostic standards, ultimately improving acceptance among clinicians.

Application of DNA methylation classification in routine diagnostics has advanced with unprecedented speed, catalyzed by the demonstrable clinical value of refined tumor subtyping. This rapid adoption, however, can feel challenging for users new to high-dimensional molecular diagnostics, raising questions regarding robust quality control and cross-validation of results. Although multiple classifier tools may emerge - possibly using divergent statistical strategies - the potential redundancy could bolster procedural safety^48^. At the same time, discordant outcomes among classifiers may generate confusion, particularly if each tool adopts slightly different nomenclature or thresholds for confidence scores. Clear consensus, transparent communication, and adherence to regulatory guidelines will be essential for ensuring that the field continues to move toward improved and harmonized, rather than fragmented, diagnostic utility. In this regard, the updated Heidelberg Methylation Classifier underscores both the promise and the logistical complexity of translating cutting-edge research into clinically actionable information. Ultimately, ongoing collaboration among researchers, clinicians, regulatory bodies, and commercial partners will be crucial for realizing the full potential of epigenomic classification to improve patient care in neuro-oncology.

## Methods

### DNA-Methylation Array Processing

The Illumina Infinium HumanMethylation450 (450k) array, Illumina Infinium MethylationEPIC (EPIC) array and Illumina Infinium MethylationEPICv2 (EPICv2) were used to obtain genome-wide DNA methylation data for tumor samples and normal control tissues according to the manufacturer’s instructions (Illumina, San Diego, USA). Data not gathered through molecularneuropathology.org, were generated at the Genomics and Proteomics Core Facility of the German Cancer Research Center (DKFZ, Heidelberg, Germany) and processed accordingly. DNA methylation data were generated from both fresh-frozen and formalin-fixed paraffin-embedded (FFPE) tissue samples. For most fresh-frozen samples, >500 ng of DNA was used as input material, while 250 ng of DNA was used for most FFPE tissues. On-chip quality metrics of all samples were carefully controlled.

All computational analyses were performed in R version 4.3.3 (R Development Core Team, 2024). Raw signal intensities were obtained from IDAT files using the minfi Bioconductor package version 1.21.4 (Aryee et al., 2014). Illumina EPIC, EPICv2 and 450k samples were merged into a combined dataset by selecting the intersection of probes present on both arrays (combineArrays function in minfi). Each sample was individually normalized by performing a background correction (shifting the 5^th^ percentile of negative control probe intensities to 0) and a dye-bias correction (scaling of the mean of normalization control probe intensities to 10,000) for both color channels. Subsequently, a correction for the type of material (FFPE/frozen) and array type (450k/EPIC(v2)) was performed by fitting univariable linear models to the log2-transformed intensity values (removeBatchEffect function in the limma package version 3.30.11). The methylated and unmethylated signals were corrected individually. Beta-values were calculated from the retransformed intensities using an offset of 100 (as recommended by Illumina).

### Non-linear dimension reduction

To perform unsupervised non-linear dimension, the 10,000 CpG probes with the highest standard deviation were selected, and a UMAP projection was calculated using the umap function available in the R-package uwot (https://github.com/jlmelville/uwot).

### Classifier training

Classifier training was performed as described in Capper et al. (2018)^1^ and Maros et al. (2020)^43^. First, we applied a permutation-based variable importance measure (R-package randomForest v4.7-1.2) to select the 10,000 most informative CpG probes as features for the final Random Forest (RF). Unbalanced class sample sizes were taken into account by down sampling each bootstrap sample to the minority class. Next, a ridge-penalized multinomial logistic-regression model (R-package glmnet v4.1-8) was fitted to calibrate the RF output, mapping raw prediction scores to probability estimates. An optimal penalization parameter was chosen by a ten-fold cross-validation. Combining classifier outputs with a logistic regression model is an ensemble strategy known as stacking^49^.

### Classifier validation

To evaluate the classifier, a five-fold nested cross-validation scheme generated out-of-sample RF scores that enabled us to fit and validate the calibration models in each fold. To measure the performance of the classifier the following metrics and figures were generated: Accuracy, Balanced Accuracy, F1, Matthews Correlation Coefficient (R-package mltest v1.0.1), Confusion Matrix, multiclass Log Loss, multiclass Brier Score, Calibration Plots (R-package rms v6.8-2). In addition, receiver operating characteristics (ROC) curves and accompanying areas under the curve (AUC) are generated using R-package pROC v1.18.5.

## Data sharing

The training dataset has been deposited in the German Human Genome-Phenome Archive (GHGA) under reference number GHGAS89861553411214.

## Declaration of generative AI and AI-assisted technologies in the writing process

During the preparation of this work the authors used ChatGPT (OpenAI) in order to refine language, enhance clarity and coherence, and improve overall readability of the manuscript. After using this tool/service, the authors reviewed and edited the content as needed and take full responsibility for the content of the publication.

## Data Availability

The training dataset has been deposited in the German Human Genome‐Phenome Archive (GHGA) under reference number GHGAS89861553411214.

https://data.ghga.de/browse?q=GHGAS89861553411214

